# Azithromycin in patients with Covid-19; a systematic review and metanalysis

**DOI:** 10.1101/2021.08.03.21261414

**Authors:** Luis Ayerbe, Ivo Forgone, Carlos Risco-Risco, Maria Pérez-Piñar, Salma Ayis

## Abstract

**Background:** Azithromycin (AZM) has been widely used in the management of Covid-19. However, the evidence on its actual effects remains disperse and difficult to apply in clinical settings. This systematic review and metanalysis summarizes the available evidence to date on the beneficial and adverse effect of AZM in patients with Covid-19.

**Methods:** The PRISMA 2020 statement criteria were followed. Randomized controlled trials (RCTs) and observational studies comparing clinical outcomes of patients treated, and not treated, with AZM, indexed until the 5^th^ of July 2021, were searched in PubMed, Embase, The Web of Science, Scopus, The Cochrane Central Register of Controlled Trials, and MedRXivs. We used Random-effects models to estimate pooled effect size from aggregate data.

**Results:** The initial search produced 4950 results. Finally, 16 studies, five RCTs and 11 with an observational design, with a total of 22984 patients, were included. The metanalysis showed no difference in mortality for those treated, or not, with AZM, OR: 0.95 (0.79-1.13). There was also no significant difference for those treated, and not, with AZM in need for hospital admission or time to admission from ambulatory settings, clinical severity, need for intensive care, or adverse effects.

**Conclusions:** These results presented in this review do not support the use of AZM in the management of Covid-19. They also show that any harm caused to the patient who received it is unlikely. Future research on treatment for patients with Covid-19 may need to focus on other drugs.

## Introduction

Azithromycin (AZM) has been widely used in the management of Covid-19.^1,2^ It is a broad-spectrum antibiotic, which is rapidly absorbed after oral intake, and has a long half-life. Evidence suggests that AZM has antiviral activity in bronchial epithelial cells, together with anti-inflammatory and immunoregulatory effects.^1,2^ The association of AZM with improved outcome in patients with other viral pneumonias, and in those with acute lung injury admitted in intensive care, has also been reported.^2^ The possible beneficial effect of AZM in patients with Covid-19 and bacterial superinfection has been considered as well.^1^ It is an economical drug that can be used in early stages of Covid-19. However, the possible QT-prolongation and cardiotoxicity associated with AZM are a concern.^1-4^

A number of individual studies have investigated the effect of AZM on different clinical outcomes among patients with Covid-19. The reviews where these articles are summarized are either narrative, they focus on the effect of AZM in combination with hydroxychloroquine, or are restricted to studies with a particular design. Therefore, the evidence on the actual beneficial or harmful effect of AZM in patients with Covid-19 remains disperse and difficult to apply in clinical settings.^1^ This systematic review and metanalysis summarizes the evidence on the beneficial and adverse effect of AZM in patients with Covid-19.

## Methods

This systematic review and metanalysis was registered in the International prospective register of systematic reviews (PROSPERO) with the reference CRD 42021252219, and it was conducted following the PRISMA 2020 statement criteria.^5,6^ Randomized controlled trials and observational studies comparing clinical outcomes of patients treated, and not treated, with AZM, were searched and considered for inclusion. All the publications indexed up to the 5^th^ of July 2021, in the following six databases were reviewed: PubMed, Embase, The Web of Science, Scopus, The Cochrane Central Register of Controlled Trials (CENTRAL), and MedRXivs. The following search strategy was used: ((“Azithromycin”[Mesh]) OR “Macrolides”[Mesh]) AND (((“Coronavirus”[Mesh]) OR “COVID-19”[Mesh]) OR “SARS-CoV-2”[Mesh]) Studies in the bibliography of all the relevant reviews identified in the initial search were also considered for inclusion. The forward citation tool in the Web of Science was used and all papers that cited those included in the review were also considered for inclusion. There were no restrictions on the basis of language, sample size or duration of follow-up. Studies were not included in the following cases: they reported outcomes of specific participants i.e: cancer patients only; there was no comparison arm; azithromycin was compared against an intervention different to placebo or standard care; the effect of azithromycin in combination with another drug was the objective of the study; the exposure was not specifically azithromycin i.e: antibiotics, macrolides; the study had an observational design but no multivariate analysis, with adjustment for potential confounders, had been conducted.

Where several articles reported results from the same population, data were taken from the publication with the longest follow-up. The quality of all studies was assessed according to accepted criteria.^7^ Authors of studies were contacted when it was unclear whether papers met the inclusion criteria, and to verify methods and results that may not have been reported. We used Random-effects models to estimate pooled effect size from aggregate data.^8^ The majority of the studies, provided estimates on the risk of death, Odds Ratios (ORs), Relative Risk (RRs), or Hazard Ratios (HRs). We pooled these using ORs, where RRs were transformed to ORs, and HRs were used as proxy measures for ORs as the percentage of the outcome was relatively small and the follow up period was short.^9^ When an RCT did not report a summary estimate, but provided numbers of deaths in each exposure group, the OR was calculated and used in the pooled summary estimates. When observational studies did not provide numerical measures of effect for death, they could not be included in the metanalysis and their results are presented narratively. Observational studies, and RCTs were displayed in one forest plot stratified by study design.

The summaries for outcomes other than death were reported narratively, due to the different methods used across studies and the small number of studies that investigated each outcome. Between-study heterogeneity was assessed using I^2^ statistics, which describes the percentage of variation across studies that is due to heterogeneity rather than chance.^11^ Publication bias was assessed visually using funnel plots and the Egger test was used to measure the small study effect. Statistical analysis was performed using the software STATA V.16.

## Results

The initial search produced 4950 results, seven of them were reviews relevant to this topic.^2,12-17^ The full text of 35 articles was assessed. Finally, our review included 16 studies, with a total of 22984 patients.^18-33^ The studies assessed in each stage of the search are presented in supplement one. Five of the studies were randomized controlled trials and 11 had an observational design. They had been conducted in Brazil, France, Italy, Iran, Spain, Turkey, the UK, and the USA. Four studies had been conducted in ambulatory settings, 11 in hospitals, and one included both hospital and ambulatory patients. The sample size ranged from 111 to 7763.^18,19^ patients. The characteristics of the studies included in this review are presented in tables one and two.

The dose of azithromycin was reported in 11 studies. In four of them patients received 500 mg a day for 5 days.^19-22^ Patients were given 500 mg on the first day, and then 250 mg a day the following four days, in three studies.^23-25^ In two studies patients were treated with 500 mg a day for ten days.^18,26^ In one study patients received 500 mg a day for 14 days^27^ and in another one 500 mg a day for 3 days.^28^

Death was the outcome in 13 studies, duration of hospital admission in six, need for intensive care unit (ICU) in four, and need for admission in two. Other secondary outcomes were also reported in individual studies. Tables one and two present the outcomes reported in each study. The quality of the studies was considered good in 15 studies, and limited in one where only an abstract was available. (Supplements 2 and 3)

The metanalysis showed no difference in death for those treated, or not, with AZM, in observational studies OR: 0.90 (0.66-1.24), RCTs, OR: 0.97 (0.87-1.08), and when studies with both designs were pooled together OR: 0.95 (0.79-1.13). (Figure 1) In the study by Hinks and colleagues^27^ the outcome was death or need for admission, and this was used as a proxy for death in the metanalysis. Excluding this study had minimal effect on the magnitude and the significance of the results, with an overall OR: 0.95 (0.79-1.14). Two further observational studies that did not present measure of effect, and could not be included in the metanalysis, reported narratively no evidence of association between treatment with AZM and death.^24,29^ Three observational studies and one RCT reported no association between the use of AZM and the duration of hospital admission,^20,24,26,31^ while another RCT showed evidence of hospital admission shorter by one day for those who received AZM.^19^ The two observational studies and the two RCTs, that had need for ICU as an outcome, reported it not to be associated with AZM.^18,19,20,29^ One observational study and one RCT, conducted both in the community, reported no association between treatment with AZM and need for admission.^22,28^

**Figure 1.**
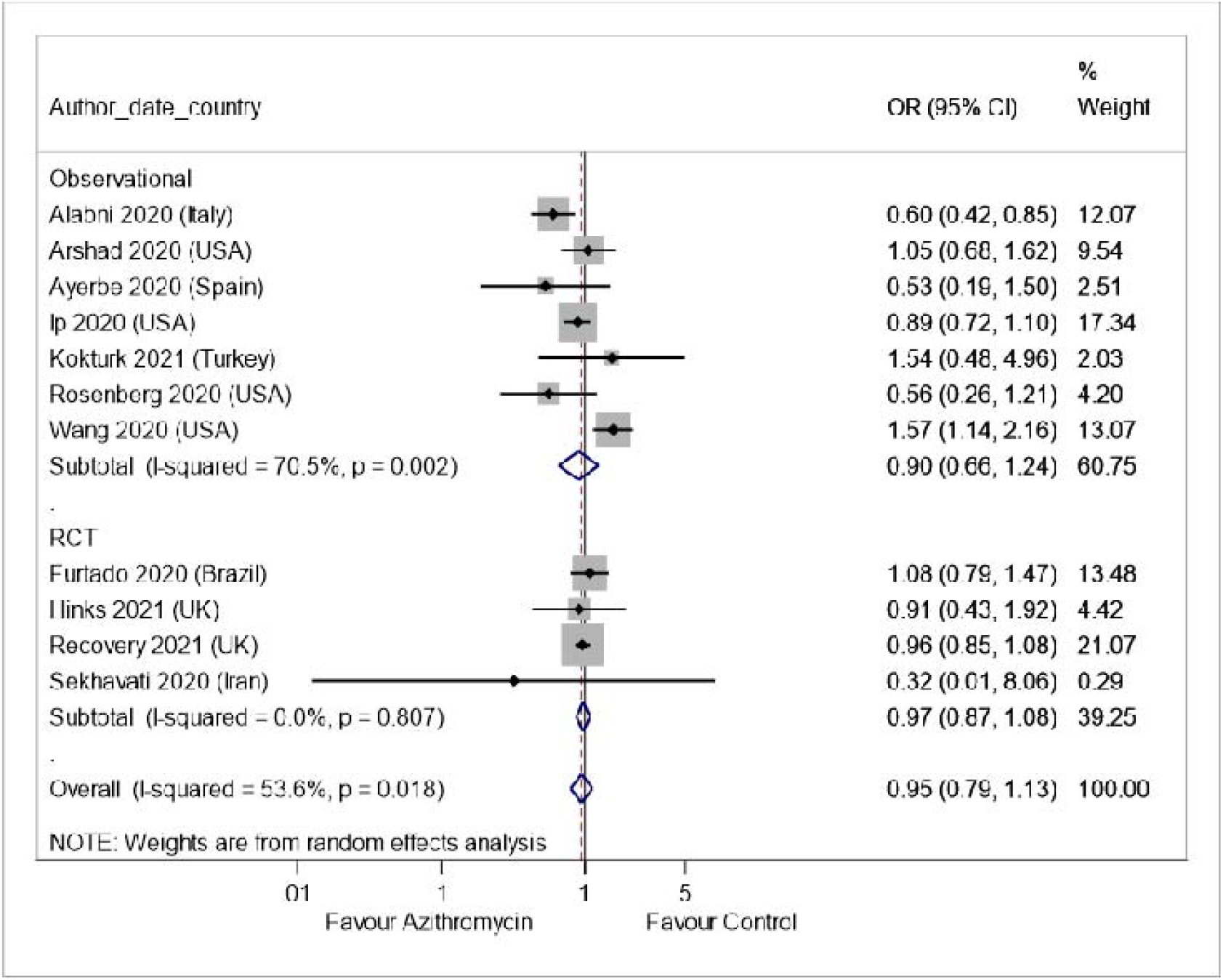
Forest plot of Observational studies and RCTs on the association between treatment with AZM and death

AZM was not associated with serious adverse events,^18,26^ QT prolongation,^19,26,30^arrythmia,^18,19,26^or need for resuscitation.^26,30^ (Tables 1 and 2) The reasonably symmetrical funnel plot supports the theory that there is no publication bias. (Supplement 4)

**Table 1.** Randomised Controled Trials included in the review (Hosp: Hospital, Comm: Community)

| Author.<br>Country | Setting | N<br>AZM/comparison | Outcome | Outcome n<br>AZM/Comparison | Effect |
| --- | --- | --- | --- | --- | --- |
| Furtado –<br>Brazil<br>2020 | Hosp | 214/183 | Higher category<br>of clinical status<br>score |  | Clinical status: 1.36(0.94-1.97) |
|  |  |  | Death | 90/73 | Death 1.08 (0.79-1.47)<br>p=0.63 |
|  |  |  | Median duration<br>of admission<br>(days) | 26/18 | Median Difference 8.00<br>(0.81 -15.19) p=0.064 |
|  |  |  | Serious adverse<br>events | 102/75 | No difference in adverse<br>events p=0.35 |
|  |  |  | QT prolongation | 47/42 | No difference.<br>No difference in ventricular<br>arrhythmia or need for<br>resuscitation |
| Hinks<br>2021 UK | Comm | 147/148 | Death or need for<br>admission | 15/17 | OR: 0.91(0.43-1.92) p=0.80 |
|  |  |  | Time to<br>admission |  | HR: 0.95(0.46-1.96)p=0.89 |
|  |  |  | Maximum<br>clinical severity |  | OR: 0.91(0.57-1.46) p=0.69 |
| PRINCIPLE 2021<br>UK | Comm | 540/875 | Clinical recovery<br>day 28 | 402/631 |  |
|  |  |  | Time to clinical<br>recovery |  | HR: 1.08(0.95-1.23) |
|  |  |  | Need for<br>Admission | 16/28 | Difference: 0.3(-1.7-2.2) |
| RECOVERY<br>UK<br>2021 | Hosp | 2582/5181 | Death | 561/1162 | RR: 0.97 (0.87-1.107)<br>p=0.50<br>This result was also not<br>significant in different age<br>or gender categories |
|  |  |  | Median duration<br>of admission<br>(days) | 10/11 |  |
|  |  |  | Discharged alive<br>on day 28 (%) | 68/69 | RR: 1.04(0.98-1.10)p=0.19 |
|  |  |  | Need for<br>mechanical<br>ventilation or<br>death (%) | 25/26 | RR: 0.95(0.87-1.03)p=0.24 |
|  |  |  | Serious adverse<br>events | 1/0 | No difference in arrhythmias |
| Sekhavati<br>Iran<br>2020 | Hosp | 56/55 | Duration of<br>admission<br>(Mean?days) | 4.61/5.96 | P=0.02 |
|  |  |  | Need for ICU | 2/7 | P=0.070 |
|  |  |  | Death | 0/1 | p=0.495 |
|  |  |  |  |  | No difference in QT<br>prolongation or arrhythmia |

**Table 2.** Observational studies included in the review (Hosp: Hospital, Comm: Community)

| Author<br>year<br>Country | Setting | N<br>AZM/Comparison | Outcome | Outcome n<br>AZM/Comparison | Effect |
| --- | --- | --- | --- | --- | --- |
| Albani<br>2020<br>Italy | Hosp | 421/605 | Death | 69/172 | OR 0.60 (0.42–0.85) |
|  |  |  | Need for ICU | 20/46 | OR: 1.08(0.57-2.05) |
|  |  |  | Median Duration of admission (days) | 6/6 | OR: 1.17(1.10-1.25) |
| Arshad<br>2020 USA | Hosp | 147/409 | Death | 33/108 | HR: 1.050 (0.682-1.616) |
| Ayerbe<br>2020<br>Spain | Hosp | 1223/796 | Death | 146/140 | OR: 0.53(0.19-1.50)p=0.233 |
| Guérin<br>2020<br>France | Comm | 34/34 | Time to clinical recovery (days) | 12.9/25.8 | (P=0.0149)<br>No serious adverse event<br>neither cardiovascular<br>events were reported in any<br>treatment group. |
|  |  |  | Need for ICU | 8/1 |  |
| Ip<br>2020<br>USA | Hosp | 256/2256 | Death |  | HR: 0.89(0.72-1.10) p=0.28 |
|  |  |  | Duration of admission |  | HR: 1.45 (0.88-2.41)p=0.150 |
| Kokturk<br>2021<br>Turkey | Hosp | 738/762 | Death | 34/33 | OR: 1.54 (0.48-4.98) p=0.472 |
| Pathak<br>2021 USA | Hosp |  | Need for ICU<br>ICU death |  | No association between AZM<br>and the outcomes |
| Rodriguez<br>-Molinero<br>2020<br>Spain | Hosp | 120/63 | Death | 7/6 | P=0.501 |
|  |  |  | Duration of admission |  | HR: 1.45 (0.88-2.41)p=0.150 |
| Rosenberg<br>2020<br>USA | Hosp | 121/211 | Death | 21/28 | HR: 0.56(0.26-1.21) |
|  |  |  | Cardiac arrest | 5/7 | HR: 0.64(0.27-1.56) |
|  |  |  | Abnormal ECG |  | HR: 0.95(0.47-1.94) |
| Szente-Fonseca<br>2020<br>Brazil | Comm | 380/337 | Need for Admissions (114) |  | 0.93(0.60-1.45) |
| Wang<br>2020<br>USA | Hosp and comm | 535/4165 | Death | 124/168 | OR: 1.57(1.14-2.16) p=0.006 |

## Discussion

This systematic review and metanalysis presents strong evidence on the lack of association between AZM and any clinical benefit.^34^ This evidence is consistent across 16 studies conducted in Europe, America and Asia with diverse methodology and design. There is also no evidence of AZM being associated with any serious adverse events, including cardiovascular disease.

This review has some limitations. Only one person extracted most of the data (LA). Even so, all data were checked for accuracy on repeated occasions and all analyses were conducted several times and checked by a senior statistician (SA). It is also possible that some publications may have been missed. The use of standard care, provided in addition to AZM in the intervention arm, and on its own in the comparison arm, was not described in detail in some studies. It is not clear how different this standard care was across the studies and how this may have affected the results. Some RCTs adjusted the treatment effect for confounders but not all did so. In those adjusted however, there was no considerable difference between the unadjusted and adjusted estimates, likely due to the balanced characteristics of the compared groups, achieved by randomization.

The comprehensive search in six databases, and critical assessment of 16 studies, that added together a large number of patients, represent strengths of this review. The inclusion of both observational and interventional studies, based in different settings and looking at various outcomes, are also positive aspects of this research. Furthermore, the sensitivity analyses that was conducted adds consistency to the results. The use of a random effect model was a conservative choice. The overall estimate remained significant despite the increased width of the confidence intervals, providing support to the significance of the findings.

The results presented in this review do not support the use of AZM in the management of Covid-19. They also show that any harm caused to patients who received it is unlikely, which would be consistent with the well stablished safety profile reported before for AZM.^4^ Future research on repurposed or innovative treatment for patients with Covid-19 may need to consider alternative drugs.

## Data Availability

This study used data that is all publically available

## Transparency declarations

Authors have no interests to declare

## Authors contributions

LA had the original idea, that then received input from IF, CRR, MPP and SA. LA conducted the searches. IF and MPP assessed the quality of the studies. LA and SA extracted the data. SA conducted the statistical analyses. LA wrote the first draft, that was later amended with contributions and references provided by IF, CRR, MPP and SA.

