## Supplements 1-4 for "Azithromycin in patients with Covid-19; a systematic review and metanalysis"

Initial Search : 4950

(PubMed : 573; Embase: 1172; Web of Science: 648; Scopus: 2233; CENTRAL: 203; MedRXivs: 121)

Previous reviews related to this topic: 7

(488 references)

Not fitting inclusion criteria: 4916

Selected for full text assessment: 27

Selected for full text assessment: 2

Papers citing included studies:722

Selected for full text assessment:6

Total Selected for full text assessment: 35

Not fitting inclusion criteria:19

Studies included: 16

Supplement 1. Studies identified in each stage of the search

|  | Furtado | Hinks | PRINCIPLE | RECOVERY | Sekhavati |
| --- | --- | --- | --- | --- | --- |
| Appropriate and clearly focused question. | + | + | + | + | + |
| The assignment of subjects to treatment groups is randomised. | + | + | + | + | + |
| Adequate concealment method | + | + | + | + | + |
| The design keeps subjects and investigators ‘blind’ about treatment allocation. | + | + | + | + | + |
| The treatment and control groups are similar at the start of the trial. | + | + | + | + | + |
| The only difference between groups is the treatment under investigation. | + | + | + | + | + |
| All relevant outcomes are measured in a standard, valid and reliable way. | + | + | + | + | + |
| What percentage of the individuals recruited into each treatment arm of the study dropped out before the study was completed? | 0% | 0% | 0% | 0% | 0% |
| All the subjects are analysed in the groups to which they were randomly allocated | + | + | + | + | + |
| Where the study is carried out at more than one site, results are comparable for all sites. | Can’t say | + | Cant say | + | Can’t say |
| Overall quality to minimise bias? | Good | Good | Good | Good | Good |

Supplement 2. Quality assessment of Randomized controlled trials

|  | Albani | Arshad | Ayerbe | Guérin | Ip | Korturk | Pathak | R- Molinero | Rosenberg | S-Fonseca | Wang |
| --- | --- | --- | --- | --- | --- | --- | --- | --- | --- | --- | --- |
| Appropriate and clearly focused question. | + | + | + | + | + | + | + | + | + | + | + |
| Selection of subjects | + | + | + | + | + | + | + | + | + | + | + |
| The two groups are selected from populations that are comparable. | NA | NA | + | + | + | + | - | + | + | NA | + |
| The study indicates how many of the people asked to take part did so | + | + | + | + | + | + | - | + | + | + | + |
| The likelihood that some subjects might have the outcome at the time of enrolment is taken into account. | + | + | + | - | - | - | + | - | - | + | - |
| What percentage of individuals dropped out before the study was completed. | 0% | 0% | 0% | 0% | 0% | 0% |  | 0% | 0% | 0% | 0% |
| Comparison is made between full participants and those lost to follow up, by exposure status. | - | - | + | - | - | - | - | - | - | - | - |
| Outcomes clearly defined. | + | + | + | + | + | + | + | + | + | + | + |
| The assessment of outcome is made blind to exposure status | NA | + | + | + | + | - | - | - | + | + | + |
| There is some recognition that knowledge of exposure status could have influenced the assessment of outcome. | + | + | - | - | - | - | - | + | + | + | - |
| Assessment of exposure is reliable. | + | + | + | + | + | + | + | + | + | + | + |
| Method of outcome assessment is valid and reliable. | + | + | + | + | + | + | + | + | + | + | + |
| Exposure level or prognostic factor is assessed more than once. | + | + | + | + | + | + | + | + | + | + | + |
| Potential confounders are identified and taken into account. | + | + | + | - | - | + | - | - | - | + | - |
| Confidence intervals provided | + | + | + | + | + | + | + | + | + | + | + |
| Overall quality to minimise the risk of bias or confounding? | Good | Good | Good | Good | Good | Good | Limited | Good | Good | Good | Good |

Supplement 3. Assessment of Observational studies

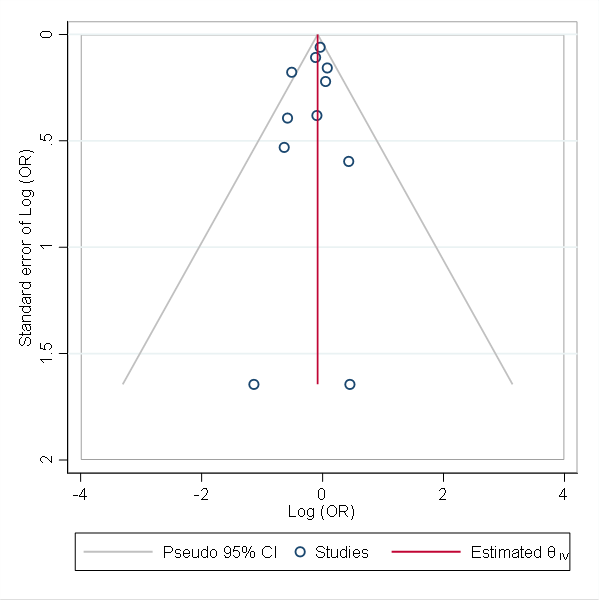

Supplement 4. Funnel plot

Egger test shows no evidence of small study effects, P Value = 0.4748
